# Real-world observation on response to cholinesterase inhibitors or selective serotonin reuptake inhibitors prescribed to outpatients with dementia using electronic medical records

**DOI:** 10.1101/2022.10.19.22281270

**Authors:** Jill S. Chotiyanonta, Kengo Onda, Milap A. Nowrangi, Xin Li, Xin Xu, Roy Adams, Constantine G. Lyketsos, Peter Zandi, Kenichi Oishi

## Abstract

**Objective:** Cholinesterase inhibitors (CEI) are prescribed for dementia to maintain or improve memory. Selective serotonin reuptake inhibitors (SSRI) are also recognized as first-line agents for psychiatric symptoms seen in dementia. What proportion of outpatients actually respond to these drugs is still unclear. Our objective was to investigate the proportion of responders to these medications in a clinical outpatient setting.

**Methods:** We used the Johns Hopkins Electronic Medical Record System to identify patients with dementia who were prescribed a CEI or SSRI for the first time between 2010 and 2021. Treatment effects were assessed through free-text entries in clinical notes documented by clinicians. Responses were scored using a simple three-point Likert scale named the NOte-based evaluation method for Treatment Efficacy (NOTE) in addition to the CIBIC-plus, a seven-point Likert scale that has been proven in clinical trials. To validate NOTE, the relationship between NOTE and CIBIC-plus, and between NOTE and change in MMSE (Mini-Mental State examination) before and after medication were examined. Inter-rater reliability was evaluated by Krippendorff’s alpha. The proportion of responders were calculated.

**Results:** The NOTE showed excellent inter-rater reliability and correlated well with CIBIC-plus and change in MMSE scores. Out of 115 CEI cases, 27.0% reported improvement and 34.8% reported stable symptoms in cognition; out of 225 SSRI cases, 69.3% reported improvement in neuropsychiatric symptoms. Adverse events were reported in 43.5% of CEI and 23.6% of SSRI cases.

**Conclusion:** NOTE showed high validity in measuring the pharmacotherapy effects based on unstructured clinical entries. Among outpatients, more than a quarter reported improvement and more than a third reported stable symptoms with CEI, which are consistent with what was reported in clinical trials for Alzheimer’s disease. Similarly, two-thirds responded to SSRI, which is also consistent with results reported in clinical trials of agitation or depression associated with Alzheimer’s disease.

**What is already known on this topic:** Clinical trials with strict inclusion and exclusion criteria have been conducted on the effectiveness of pharmacotherapy for Alzheimer’s disease, reporting the effectiveness of cholinesterase inhibitors (CEIs) for memory impairment and selective serotonin reuptake inhibitors (SSRIs) for neuropsychiatric symptoms; however, when CEIs and SSRIs are prescribed to heterogeneous patients with dementia in routine practice, it remains unclear what proportion of patients benefit from these drugs.

**What this study adds:** This retrospective analysis of electronic medical records adds real-world perspective of treatment effects of CEIs and SSRIs on dementia.

**How this study might affect research, practice or policy:** This study will contribute to clinical decision making by providing information on the response and non-response rates of CEIs and SSRIs prescribed for dementia in everyday clinical practice.

## Introduction

Dementias including Alzheimer’s disease (AD) are progressive neurodegenerative conditions that impair cognitive function, commonly present with neuropsychiatric symptoms,^1^ and eventually require individuals to be under constant care and supervision. Currently available medications to treat dementia symptoms include cholinesterase inhibitor (CEI) to address cognitive symptoms and selective serotonin reuptake inhibitor (SSRI) to address certain neuropsychiatric symptoms. These medications have been shown to have modest, but important, clinical benefits.^2-4^ However, patients presenting with cognitive and behavioral dementia symptoms respond differentially to these medications. Medication effectiveness varies among individuals^5 6^ in part because patients with cognitive decline can be highly heterogeneous in their underlying pathology.^7 8^

Although pharmacotherapeutic effects of anti-dementia drugs and antidepressants used to treat dementia symptoms have been studied under strict inclusion and exclusion criteria in clinical trials, the proportion of outpatients who actually respond to these drugs is still under investigation.

Our aim is to study the relative responder rates of CEIs and SSRIs in an outpatient setting. To this end, our study focused on utilizing electronic medical records (EMRs) which contain a massive amount of routinely collected clinical data of outpatients with dementia. Though clinical notes are rich with information, extracting medication response from EMRs remains a challenge. As part of this study, we developed a novel method for annotating clinical notes with evidence of response to medication and assess the validity and reliability of this method. Retrospective analysis of EMRs using this method adds the real-world perspective of pharmacotherapeutic effects on patients with dementia due to various pathologies.

## Methods

### Data source and cohort screening

Retrospective analysis of Johns Hopkins EMRs was performed on patients with dementia who were prescribed CEI or SSRI for the first time between 2010 and 2021. Patients of interest were identified from the Richman Family Precision Medicine Center of Excellence in Alzheimer’s Disease database through Precision Medicine Analytics Platform, which consists of clinical records of patients seen at the Johns Hopkins Bayview Memory and Alzheimer’s Treatment Center. Because this data is part of the larger dataset used for brain imaging analysis, the inclusion criteria included the following: 1) first documented order of CEI and/or SSRI as an *index date*, 2) age 50 or older at the time of index date, 3) had any encounter diagnosis of cognitive disorder, 4) had brain MRI scans within a year before and after index date, and 5) had at least one encounter within a year before and after index date. The diagnosis codes, or ICD-10, of interest included F01 (vascular dementia (VD)), F02 (dementia in other diseases classified elsewhere), F03 (unspecified dementia), G30 (AD), and G31 (other degenerative diseases of the nervous system). Additionally, the following CEIs that are unrelated to dementia treatment were excluded: neostigmine (Bloxiverz) and pyridostigmine (Mestinon).

### Manual review of EMRs

After the initial screening, patients’ clinical records were manually reviewed in descending order of patient ID, and records whose review was completed by April 2022 were included in this article. Because the screened index dates (i.e., the first prescription-ordered date) did not always match the actual medication start date reported by patients and caregivers, the following cases were excluded: individuals who 1) had started SSRI/CEI more than 6 months prior to or after 3 months from index date, 2) were on serotonin–norepinephrine reuptake inhibitor (SNRI) less than 3 months before starting SSRI, 3) had unknown medication start date, 4) had inaccessible, protected records, 5) had no evaluable follow-up, 6) reported that they never started the medication, and 7) had medication discontinued due to change in the original diagnosis. For those who were not excluded, any relevant information in a sentence or paragraph was collected into spreadsheet as raw data. Manual review was performed by a research data analyst (JSC) who has been trained under a neurologist (KOi).

Due to the nature of dementia which involves loss of insight and progressive worsening in cognitive function, clinician comments were prioritized over caregiver comments, and caregiver comments were prioritized over patient comments as available. Medication was marked as effective whenever improvement was noted even if decline in the overall patient’s condition was reported later. Furthermore, for patients who were prescribed with more than one SSRI or CEI in their clinical history, we set a washout period of 3 months to account for possible carryover effects between different medications. Medications used from the same class was considered as two separate SSRI or CEI only if the switch was made after a 3-month period.

In addition to abstracting clinical comments, age at start of medication, sex, medication start and end dates, date of treatment effect reported, any adverse event (AE) and the corresponding date were recorded. AEs included any side effects or allergic reactions that occurred during treatment that may or may not be related to medication usage; not available (N/A) was marked when there was no recording of either AE or medication tolerance. Results and dates of Mini-Mental State Examination (MMSE) and Montreal Cognitive Assessment (MoCA) were also recorded whenever possible.

### Quantifying treatment responses

To translate evidence from a clinical note into a simple, quantitative measure of medication response, we used Likert scales. We used a 3-point Likert scale named *NOte-based evaluation method for Treatment Efficacy* (NOTE), with categories 1=improved, 2=no change, and 3=worse. For comparison, we also applied a widely-used global assessment of anti-dementia drugs called the Clinician’s Interview-Based Impression of Change Plus Caregiver Input (CIBIC-plus).^9 10^ CIBIC-plus consists of 7-point Likert scale to rate the degree of change after medication usage, with 1 indicating markedly improved and 7 indicating markedly worse. We used two versions of these two Likert scales by classifying clinical comments into psychiatric and cognitive note. Cognitive-NOTE and Cognitive-CIBIC-plus rated a change in cognitive symptoms, and psychiatric-NOTE and psychiatric-CIBIC-plus rated a change in psychiatric symptoms. Comments like “anxiety better” were classified into psychiatric measure and comments like “memory worse” were classified into cognitive measure. When comments were not readily classifiable to either psychiatric or cognition, such as “sertraline working well” or “donepezil helpful,” then the psychiatric was prioritized for SSRIs and the cognitive was prioritized for CEIs. Of note, because continued decline in cognition overtime is expected, the word “stable” was evaluated differently for psychiatric versus cognitive symptoms. Cognitive comments like “dementia stable” were classified as no change while psychiatric comments like “agitation/irritation now stable” were classified as improved. Patients using CEI and SSRI at the same time were evaluated and scored separately using available cognitive and psychiatric comments. For both scales, when it was not possible to score a treatment effect, 0=N/A was marked to indicate not available or applicable. Table-1 summarizes the keywords searched and words found within EMRs that were classified for Likert scales.

**Table-1.**
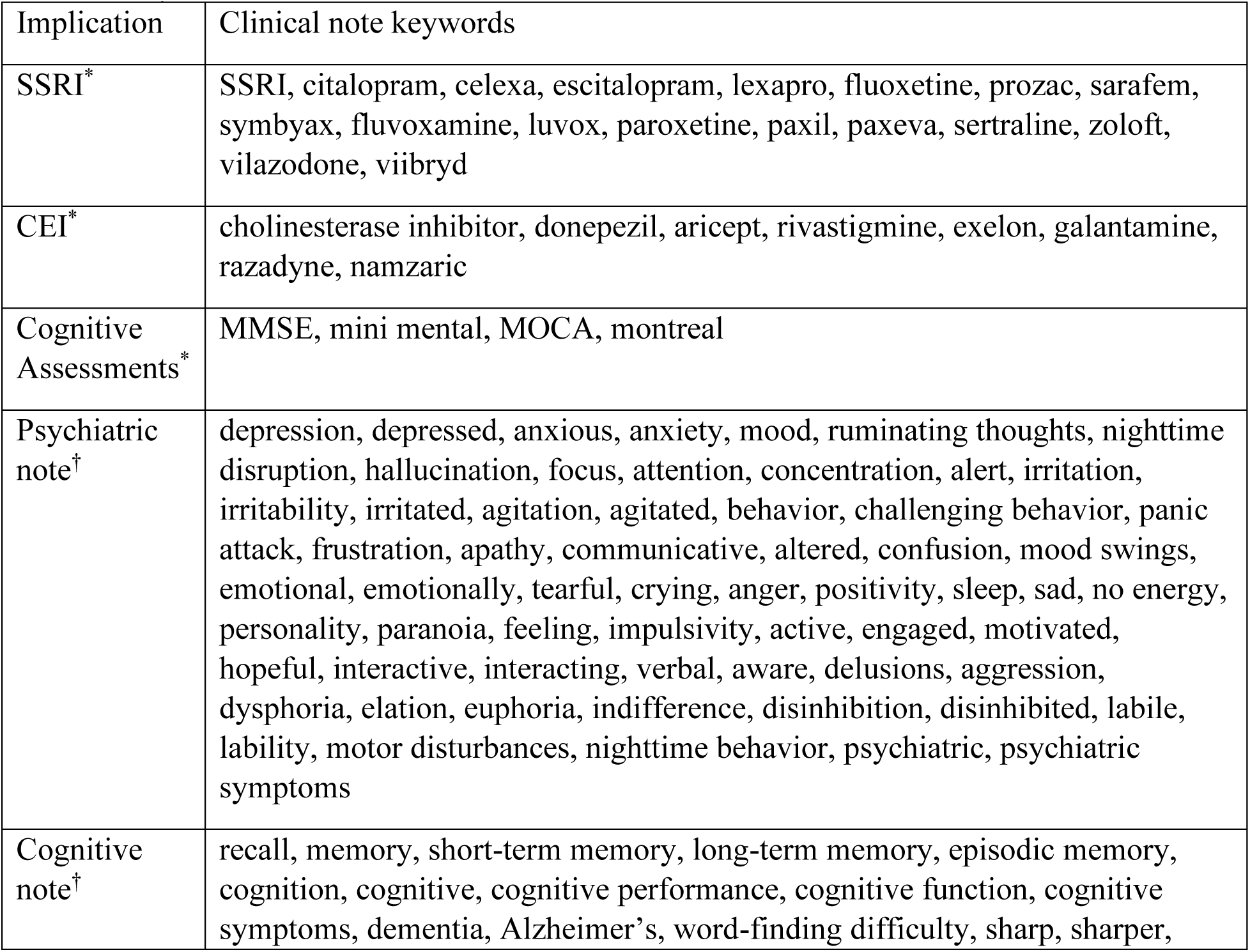
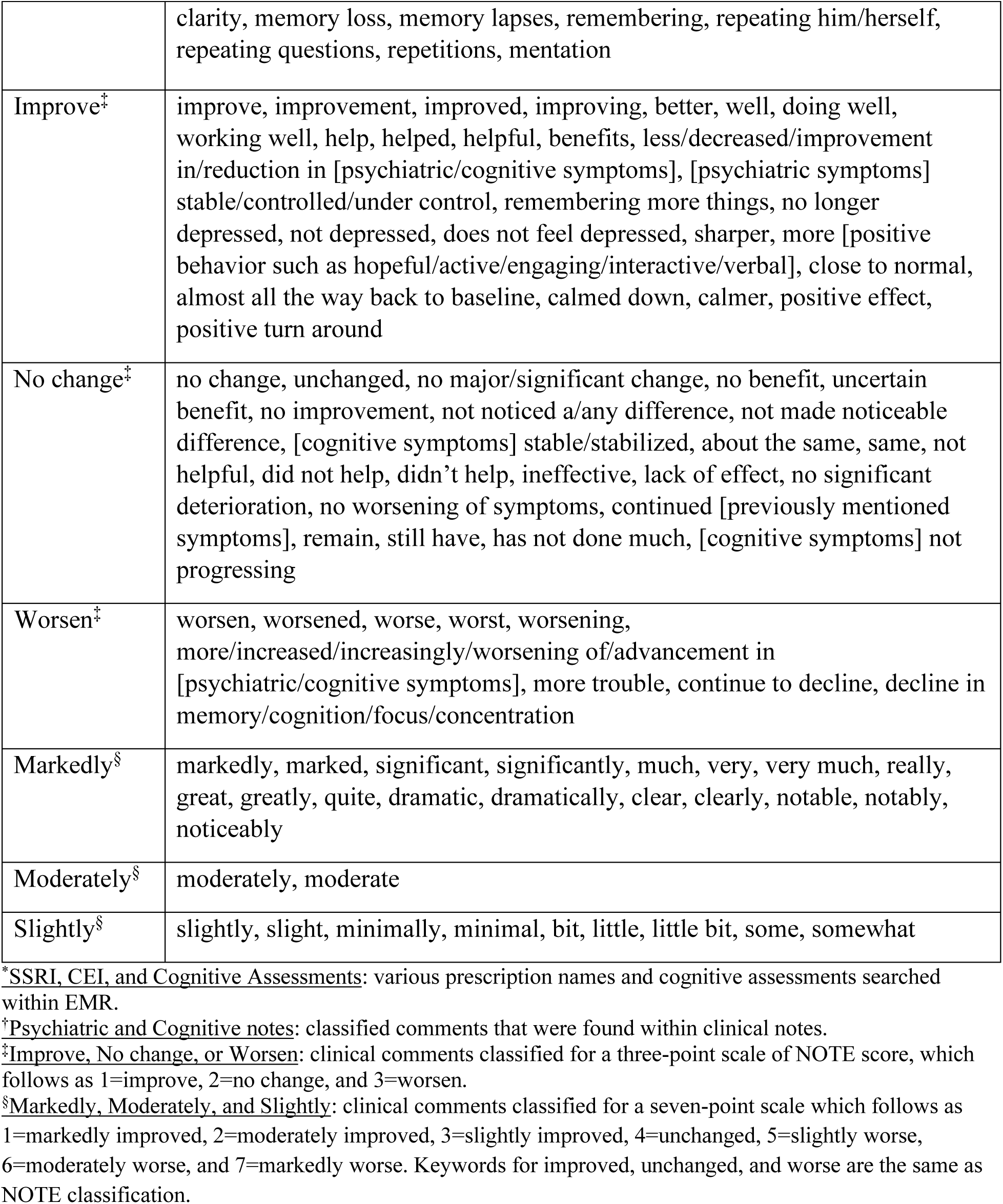
Keywords found within EMR

### Definition of responder, non-responder, and intolerance

SSRI responders were defined as psychiatric cases evaluated as improved. SSRI non-responders were defined as psychiatric cases evaluated as no change or worse. CEI responders were defined as cognitive cases evaluated as improved or stable (i.e., no change). CEI non-responders were defined as cognitive cases evaluated as worse. Intolerance was marked when clinical notes stated that individuals discontinued medication use due to AE(s) or the medication was added to allergy list; these were marked with a score of 0.5=IT for both scales.

### Statistical analyses

Inter-rater reliability of the Likert scales was evaluated by having another reviewer (KOi) collect and evaluate information on randomly selected 12 patients from EMRs. For the reliability tests, analyses of intraclass correlation coefficient (ICC) and Krippendorff’s alpha were performed on psychiatric and cognitive Likert scales of two raters using the psych (version 2.1.9)^11^ and the irr (version 0.84.1)^12^ libraries in R 4.2.0^13^, respectively. ICC of a two-way random-effects model with significance set at *P*<0.05 was calculated based on the ordinal assumption (worsen < no change < improved) by excluding N/A values. Krippendorff’s alpha-reliability was calculated to include missing data.^14^ To add these tests, dice coefficients for each NOTE were evaluated. To determine whether the NOTE introduced in this study correlated well with the established CIBIC-plus scores, the Spearman’s rank correlation test with significance set at *P*<0.05 was conducted on responses that were evaluable by both scales.

All MoCA scores were converted to MMSE scores using the MMSE-MoCA conversion table of Roalf et al.^15^ and treated as MMSE scores. MMSE change was calculated as the difference between a) the secondary MMSE score within 2 weeks of the date a cognitive clinical comment was made and b) the initial MMSE score within 2 weeks of the drug initiation. The Spearman’s rank correlation test with significance set at *P*<0.05 was also performed between NOTE and changes in MMSE from pre to post medication use. Python 3.8 was used for these statistical analyses and data presentations.

The proportion of responders, non-responders, and intolerances were analyzed in relation to age, sex, and ICD-10 codes. To see if the drug responses were affected by these characteristics, multinomial baseline-category logit model analyses were performed using the VGAM (version 1.1-5)^16^ library in R 4.2.0.

## Results

### Demographics and clinical characteristics

Initial screening identified a total of 772 patients of interest. Individuals manually reviewed for clinical notes included *n*=446 for SSRI and *n*=163 for CEI. Figure-1 summarizes the inclusion/exclusion criteria and the number of *n* for each screening stage. After excluding *n*=251 SSRI and *n*=62 CEI-users due to their medication usages and record unavailability, 195 patients on SSRI and 101 patients on CEI were included in this study. Considering patients using more than one SSRI or CEI in their clinical history, a total of 225 SSRI and 115 CEI cases of medication usage were collected from EMRs.

Figure-2 summarizes the patient demographics of age and sex and the clinical characteristics of AEs, length of time of medication usage, and diagnostic codes. Similar trends in demographics were observed: 52.0% of SSRI and 47.8% of CEI cases were female and average ages were 73 for SSRI and 74 for CEI cases. AEs were reported in 23.6% of SSRI and 43.5% of CEI cases while 57.3% of SSRI and 33.0% of CEI cases contained no record of tolerability nor AE during treatment. Analyzing diagnostic codes, SSRI cases had 32.4% AD, 18.7% VD, and 8.9% mixed dementia; likewise, CEI cases had 47.0% AD, 11.3% VD, and 11.3% mixed dementia (Fig. 2E). The types of medications used are listed in Supplemental Figure-1; the medications that were switched within 3-months washout period for various reasons, from insurance coverage change to ineffectiveness of prescription, are listed in the order of their usage. The most used drugs were sertraline for SSRI and donepezil for CEI. The time from start of medication to the assessment of treatment effect is shown in Supplemental Figure-2.

**Figure-1.**
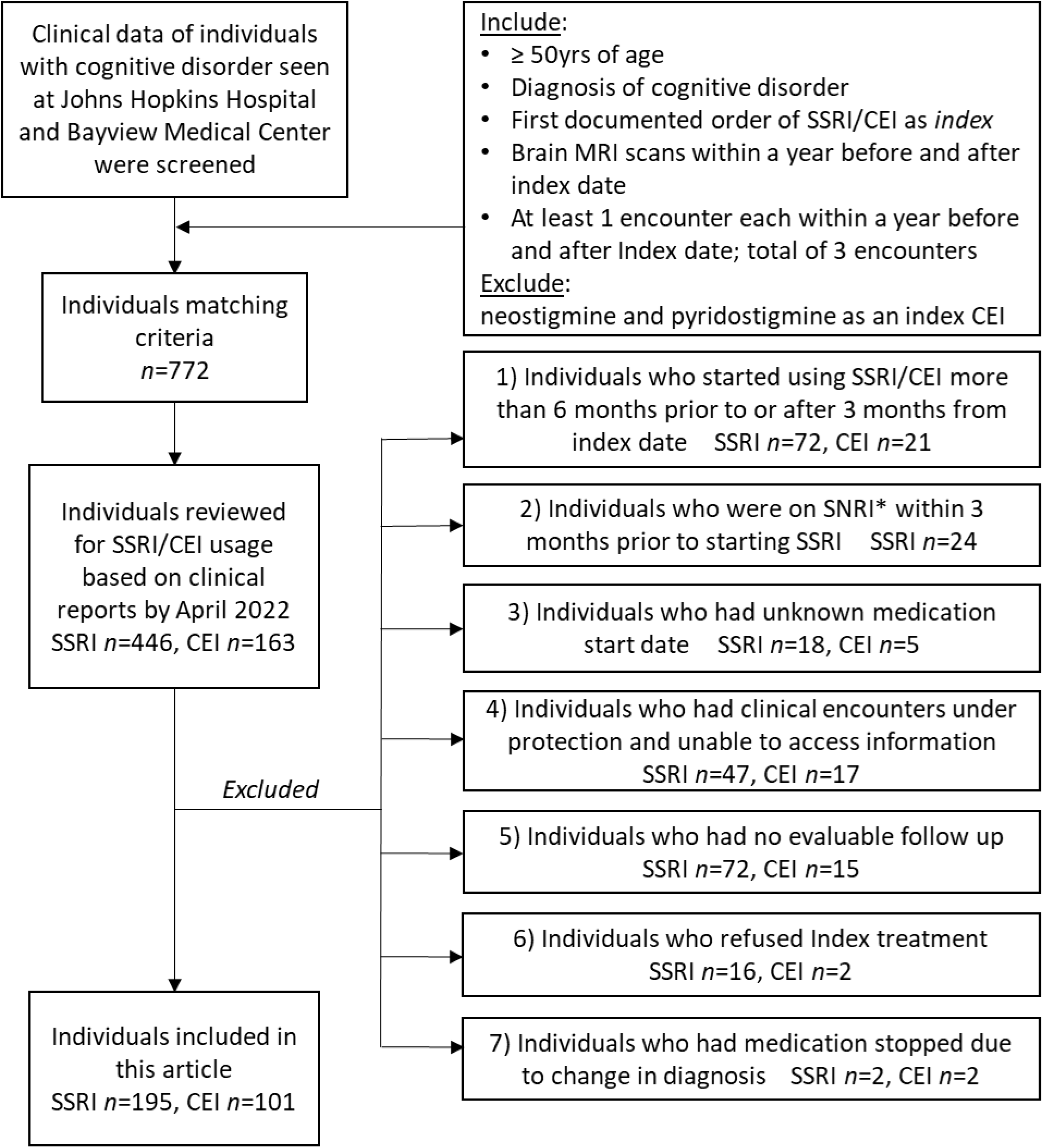
Inclusion and exclusion criteria and each *n* count. *Index* SSRI/CEI and *index date* represent the first identified prescription order and its date. The excluded cases and reasons are listed. *SNRI: serotonin–norepinephrine reuptake inhibitor

**Figure-2.**
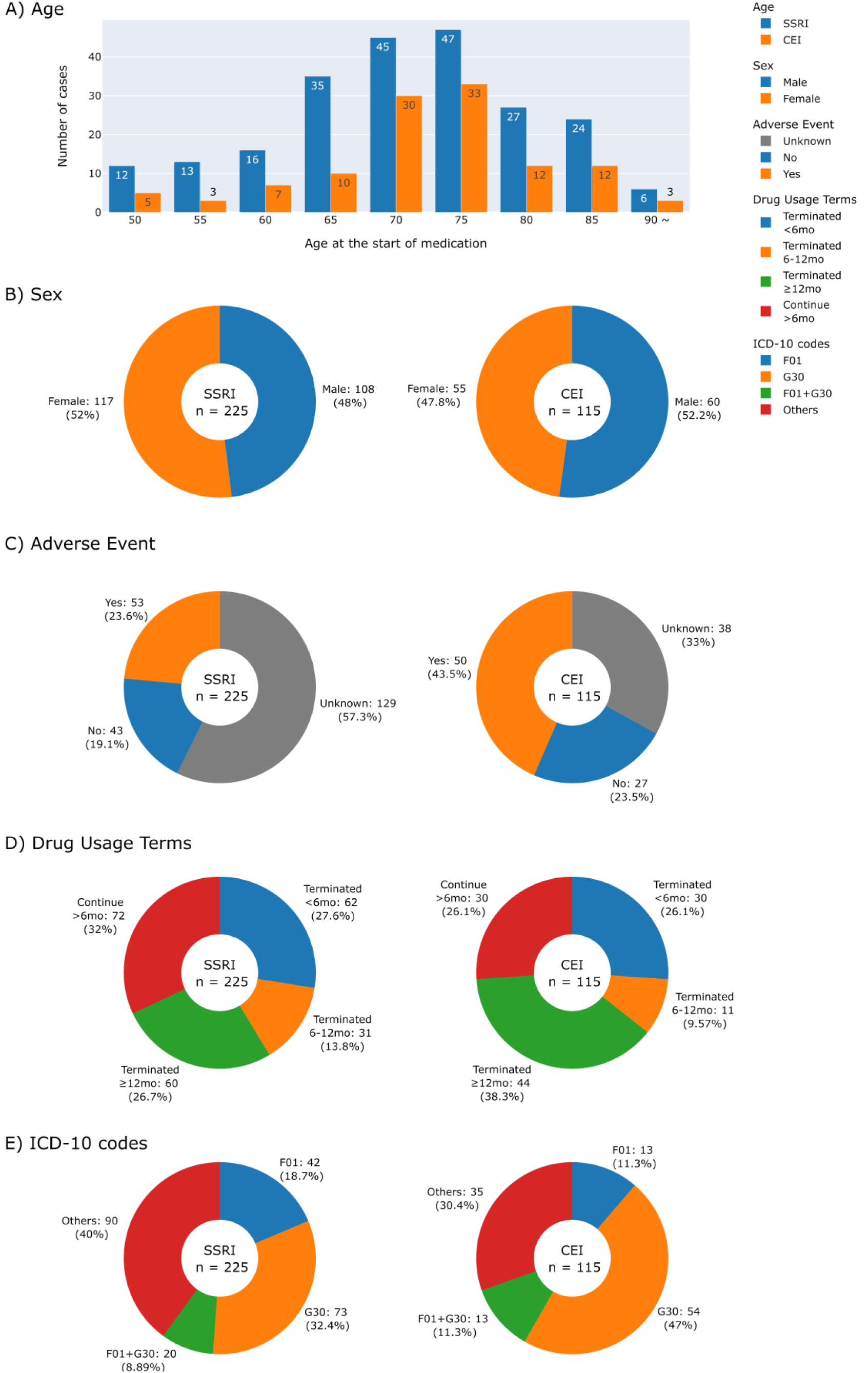
The histogram and pie charts summarizing demographics and clinical characteristics of patients by drugs. A) Age at the start of medication. B) Sex. C) Adverse Event reported. D) Drug usage terms separated into 4 groups: terminated drug use in less than 6 months, within 6 to 12 months, after 12 months, and continue for more than 6 months for those who did not have a record of terminating drug use. E) Collected ICD-10 codes were classified into F01 (vascular dementia), G30 (Alzheimer’s disease), F01 + G30 for vascular and Alzheimer’s mixed dementia, and Others for patients with other ICD-10 codes that include F02 (dementia in other diseases classified elsewhere), F03 (unspecified dementia), and G31 (other degenerative diseases of the nervous system).

### Validity of NOTE score

Inter-rater reliability test revealed excellent inter-rater agreement (Supplemental Table-1). ICC showed coefficient of 1.0 for all scales. Krippendorff’s alpha showed alpha of 1.0 for psychiatric- and cognitive-CIBIC-plus and psychiatric-NOTE, and alpha of 0.78 for cognitive-NOTE. Dice coefficients were 0.75 for “N/A” and “worse” in cognitive-NOTE, and 1.0 for all other scales.

The distributions of NOTE and CIBIC-plus scores and the number of notes deemed unevaluable are shown in Figure-3. Many of the non-prioritized evaluations were missing (SSRI 64.4% of cognitive-NOTE, CEI 49.6% of psychiatric-NOTE) because they were not the target treatment of focus; cognitive effect was not always reported after SSRI use and the same applied for psychiatric effect after CEI use. For CIBIC-plus, the keywords that were not readily classifiable into 7-point scale, such as “donepezil helping,” were marked as N/A. Similarly, comments that can be classified as “moderate” were not found and therefore marked as N/A. While only 39.6% (89/225) of SSRI and 65.2% (75/115) of CEI cases could be evaluated by their prioritized CIBIC-plus, most of SSRI (219/225, 97.3%) and CEI (108/115, 93.9%) cases could be evaluated by their prioritized NOTE. The results indicated that NOTE is more suitable than CIBIC-plus for evaluating treatment effects based on the clinical notes.

Figure 4A-B shows Spearman’s rank correlations comparing NOTE and CIBIC-plus. There were strong correlations between NOTE and CIBIC-plus for both medications (SSRI [Psychiatry]: ρ=0.93, *P*=1.6×10^−31^; CEI [Cognition]: ρ=0.99, *P*=2.7×10^−59^). Figure 4C-E shows Spearman’s rank correlations between NOTE and change in MMSE from pre- to post-medication usage. NOTE scores of SSRI, CEI, and both combined all correlated significantly with MMSE changes (SSRI+CEI [Cognition]: ρ=-0.49, *P*=0.00049, *n*=47; SSRI [Cognition]: ρ=-0.42, *P*=0.0496, *n*=22; CEI [Cognition]: ρ=-0.54, *P*=0.0054, *n*=25).

**Figure-3.**
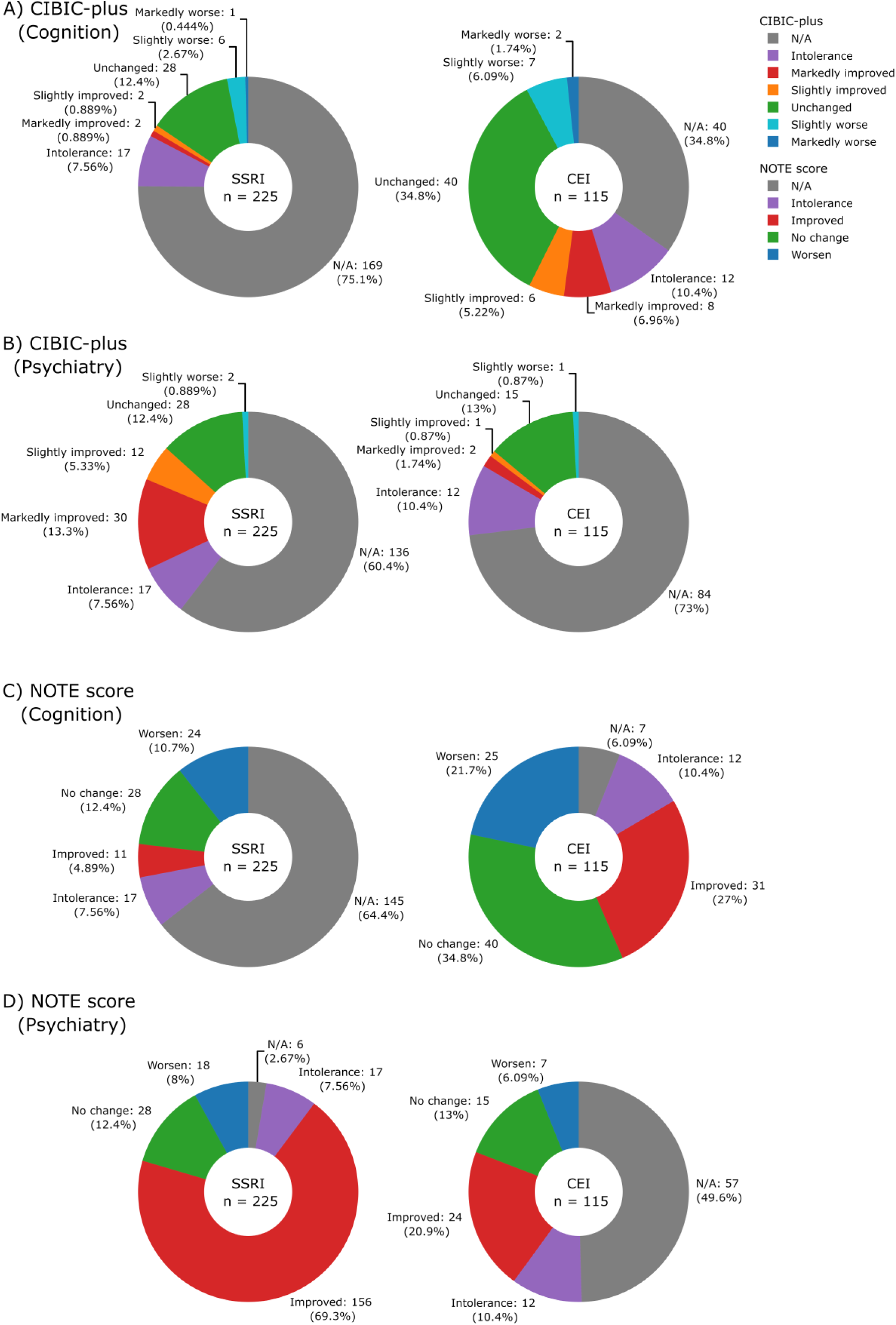
The pie charts illustrating the breakdown of CIBIC-plus (A: Cognition, B: Psychiatry) and NOTE scores (C: Cognition, D: Psychiatry) by drugs.

**Figure-4.**
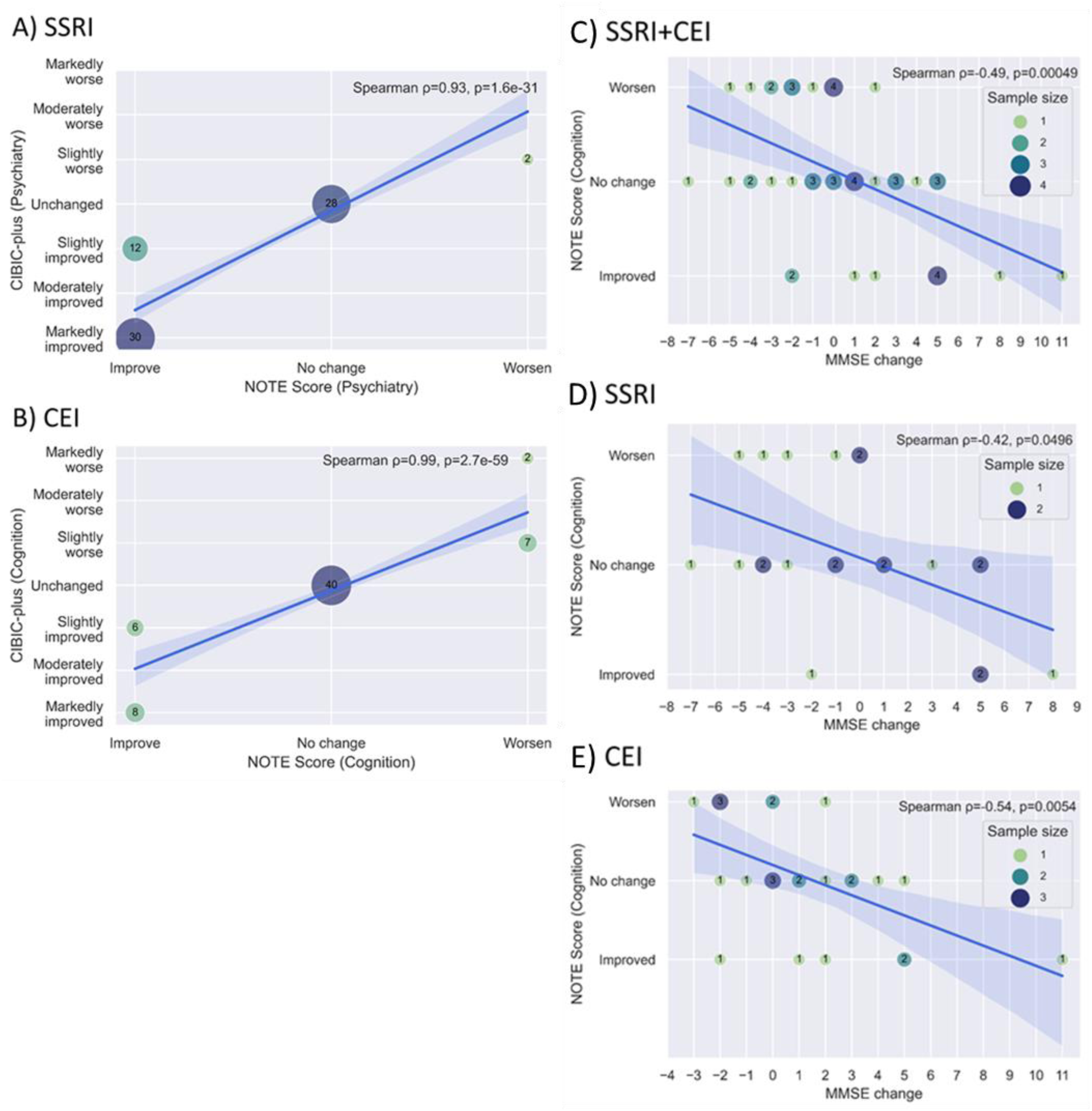
A-B) The bubble plots showing the relationships between CIBIC-plus and NOTE scores by drug (A: SSRI psychiatric scores, B: CEI cognitive scores). C-E) The bubble plots showing the relationship between MMSE changes and cognitive NOTE scores by drug (C: SSRI+CEI, D: SSRI, E: CEI). A solid blue line and a light blue area represent the regression line with its 95% confidence intervals. The coefficient and *P* value of Spearman’s rank correlation and legend of sample size (for C-E) are shown in the upper right corner of the graph.

### Treatment responses

The proportion of NOTE scores in relation to clinical characteristics are shown in Table-2. The type II likelihood ratio test on multinomial models revealed no significant differences between diagnostic codes, sex, and age in relation to the group of responders, non-responders, and intolerances among SSRI (diagnostic code: *P*=0.70; sex: *P*=0.20; age: *P*=0.45) and CEI (diagnostic code: *P*=0.66; sex: *P*=0.20; age: *P*=0.30). Among 225 SSRI cases, 69.3% were SSRI responders, 20.4% were SSRI non-responders, and 7.6% were SSRI intolerant. Among 115 CEI cases, 61.8% were CEI responders, 21.7% were CEI non-responders, and 10.4% were CEI intolerant. The remaining cases (SSRI 2.7% and CEI 6.1%) did not have the prioritized psychiatric or cognitive comment available for evaluation.

**Table-2.**
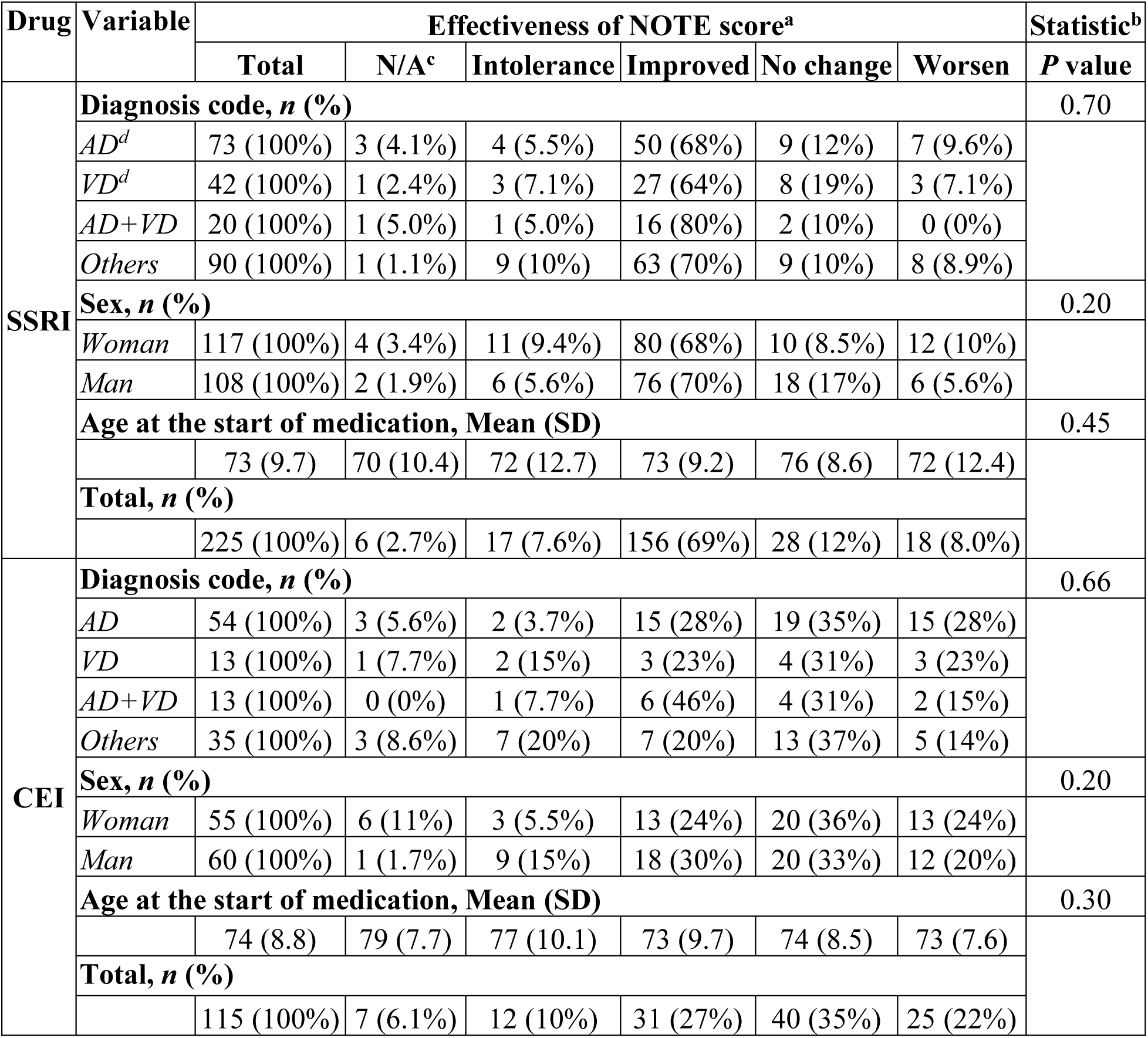
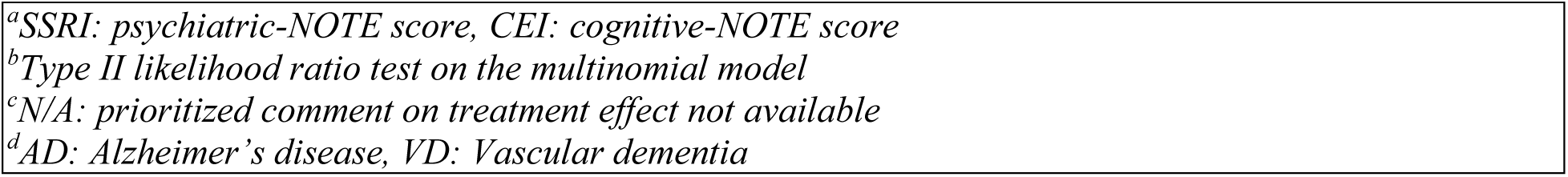
The proportion of age, sex, and diagnostic codes in relation to the results of NOTE.

| Drug | Variable | Effectiveness of NOTE score<sup>a</sup> |  |  |  |  |  | Statistic<sup>b</sup> |
| --- | --- | --- | --- | --- | --- | --- | --- | --- |
|  |  | Total | N/A<sup>c</sup> | Intolerance | Improved | No change | Worsen | *P* value |
| SSRI | Diagnosis code, *n* (%) |  |  |  |  |  |  | 0.70 |
|  | *AD*<sup>d</sup> | 73 (100%) | 3 (4.1%) | 4 (5.5%) | 50 (68%) | 9 (12%) | 7 (9.6%) |  |
|  | *VD*<sup>d</sup> | 42 (100%) | 1 (2.4%) | 3 (7.1%) | 27 (64%) | 8 (19%) | 3 (7.1%) |  |
|  | *AD+VD* | 20 (100%) | 1 (5.0%) | 1 (5.0%) | 16 (80%) | 2 (10%) | 0 (0%) |  |
|  | *Others* | 90 (100%) | 1 (1.1%) | 9 (10%) | 63 (70%) | 9 (10%) | 8 (8.9%) |  |
|  | Sex, *n* (%) |  |  |  |  |  |  | 0.20 |
|  | *Woman* | 117 (100%) | 4 (3.4%) | 11 (9.4%) | 80 (68%) | 10 (8.5%) | 12 (10%) |  |
|  | *Man* | 108 (100%) | 2 (1.9%) | 6 (5.6%) | 76 (70%) | 18 (17%) | 6 (5.6%) |  |
|  | Age at the start of medication, Mean (SD) |  |  |  |  |  |  | 0.45 |
|  |  | 73 (9.7) | 70 (10.4) | 72 (12.7) | 73 (9.2) | 76 (8.6) | 72 (12.4) |  |
|  | Total, *n* (%) |  |  |  |  |  |  |  |
|  |  | 225 (100%) | 6 (2.7%) | 17 (7.6%) | 156 (69%) | 28 (12%) | 18 (8.0%) |  |
| CEI | Diagnosis code, *n* (%) |  |  |  |  |  |  | 0.66 |
|  | *AD* | 54 (100%) | 3 (5.6%) | 2 (3.7%) | 15 (28%) | 19 (35%) | 15 (28%) |  |
|  | *VD* | 13 (100%) | 1 (7.7%) | 2 (15%) | 3 (23%) | 4 (31%) | 3 (23%) |  |
|  | *AD+VD* | 13 (100%) | 0 (0%) | 1 (7.7%) | 6 (46%) | 4 (31%) | 2 (15%) |  |
|  | *Others* | 35 (100%) | 3 (8.6%) | 7 (20%) | 7 (20%) | 13 (37%) | 5 (14%) |  |
|  | Sex, *n* (%) |  |  |  |  |  |  | 0.20 |
|  | *Woman* | 55 (100%) | 6 (11%) | 3 (5.5%) | 13 (24%) | 20 (36%) | 13 (24%) |  |
|  | *Man* | 60 (100%) | 1 (1.7%) | 9 (15%) | 18 (30%) | 20 (33%) | 12 (20%) |  |
|  | Age at the start of medication, Mean (SD) |  |  |  |  |  |  | 0.30 |
|  |  | 74 (8.8) | 79 (7.7) | 77 (10.1) | 73 (9.7) | 74 (8.5) | 73 (7.6) |  |
|  | Total, *n* (%) |  |  |  |  |  |  |  |
|  |  | 115 (100%) | 7 (6.1%) | 12 (10%) | 31 (27%) | 40 (35%) | 25 (22%) |  |

<sup>a</sup>SSRI: psychiatric-NOTE score, CEI: cognitive-NOTE score <sup>b</sup>Type II likelihood ratio test on the multinomial model <sup>c</sup>N/A: prioritized comment on treatment effect not available <sup>d</sup>AD: Alzheimer's disease, VD: Vascular dementia

## Discussion

This study aimed to find the medication responder rates of dementia patients in real-world setting. Even though our outpatient cohort included various etiologies of dementia and had no clinical exclusion criteria other than age, response rates to CEIs and SSRIs were remarkably similar to those obtained in a study cohort with a homogeneous clinical profile targeting a specific type of dementia.

In our study, 27.0% of CEI-users reported improvement and 34.8% reported stable symptoms, totaling the CEI responders to 61.8%. This is consistent with what were reported in research cohorts. According to Miranda et al., 27.8% of CEI-users were good responders scoring ≥2 in MMSE and 37.1% were neutral responders scoring between −1 and +1 in MMSE at 12 months of treatment, a total of 64.9% CEI responders.^17^ Another study reported their donepezil responder rate in AD patients as 60%.^18^ For SSRI, our study had 69.3% of outpatients reported improvement. This is also consistent with what were reported in clinical trials of agitation or depression associated with AD. According to Porsteinsson et al., 14% citalopram-users reported marked improvement, 26% reported moderate improvement, and 29% reported minimal improvement, a total of 69% citalopram responders among AD patients.^19^ Likewise, Finkel et al. reported that among dementia patients with moderate-to-severe symptoms who are on donepezil, 60% of sertraline-users achieved a response of ≥50% reduction in a four-item Neuropsychiatric Inventory-behavioral subscale.^20^

The intolerance rate was also similar to the reports based on clinical trials. While our intolerants included 7.6% of SSRI-users and 10.4% of CEI-users, a systematic review of antidepressants reported that 12% of SSRI-users withdrew from trial due to AEs,^4^ and another clinical trial reported that 10.7% of donepezil-users and 21.8% of rivastigmine-users discontinued study due to AEs.^21^

Interestingly, the proportion of diagnoses (i.e., AD vs. VD vs. AD+VD vs. other dementias), sex, and age did not differ among intolerants, responders, and non-responders, suggesting that dementia type, sex and age may not predict response to CEIs or SSRIs in heterogeneous clinical setting.

A unique quality of this study is its utilization of EMRs and analysis of natural language to assess pharmacotherapeutic responses using a simple Likert scale. Clinical trials investigating treatment effects of dementia drugs have widely used CIBIC-plus assessment in the past.^10^ CIBIC-plus has the advantage in detailing the degree of change by having a clinician interview patients and caregivers.^10 22^ However, the degree of detail necessary to translate a clinical note into a CIBIC-plus score is rarely available. Thus, in studies that involve retrospective analysis of EMRs *without* additional patient contact, NOTE provides a more flexible and effective measure. Sophisticated scales have been used in trials to assess the cognitive and functional status of dementia patients, including The Alzheimer’s Disease Assessment Scale-Cognitive Subscale^23^ and the Bristol Activities of Daily Living Scale;^22^ nonetheless these are not routinely used in clinical practice. Although the Cornell Scale and the Neuropsychiatric Inventory Questionnaire are used in clinical practice to assess neuropsychiatric symptoms, these scales are not always systematically obtained and recorded in the EMRs, making it impractical to analyze neuropsychiatric changes after medication use.

### Limitations

This study includes several limitations. First, the manual review of EMRs was labor intensive in identifying and interpreting the information of interest, which limited the study sample size. Also, because this was a retrospective study, the dates the clinical comments on treatment effect found varied widely. Moreover, with studies that involve human reviewers’ interpretations of documents, there are concerns for subjectivity and human error^24^ despite the NOTE demonstrating excellent inter-rater reliability. We expect these limitations can be addressed by incorporating artificial intelligence. The manual review presented in this study can be an important first step that can be utilized to develop a reference standard used to build Natural Language Processing (NLP) model for structuring clinical data of dementia patients for further automated, large-scale analyses of EMRs.^25 26^ Second, our analysis was performed in the EMRs with American English. Clinical text may naturally and structurally differ between healthcare systems, so the English words found and categorized in our study may not be fully generalizable to EMRs of other healthcare systems; adjustments may be necessary if similar analyses were to be conducted at other sites. Finally, the observed response rates are a product of who is prescribed a medication. For example, if clinicians are more likely to prescribe medications to patients they believe to be high-responders, then response rates may be higher than those observed if treatments were randomly assigned to a heterogeneous population. Despite this, it remains critical to know how effective a medication is in the population receiving that medication.

### Conclusion

Responses to CEIs and SSRIs prescribed to outpatients were similar to those previously reported in clinical trials. Our results also indicated that medication responses are difficult to predict by the type of dementia, sex, and age of patient. The 3-point Likert scale NOTE introduced in this study is suitable for extracting responses to drugs recorded in EMRs and has the potential to be used to train NLP models for large-scale automated analysis in the future.

## Data Availability

All data produced in the present work are contained in the manuscript.

## Acknowledgments and affiliations

We thank Dr. Brian Caffo for statistical advice. This study is supported by grants from the National Institutes of Health [R21AG070404]; and the Richman Family Precision Medicine Center of Excellence, Baltimore, MD [Venture Discovery Fund]. KOi is a consultant for “AnatomyWorks” and “Corporate-M.” This arrangement is being managed by the Johns Hopkins University in accordance with its conflict-of-interest policies.

## Ethics statements

The study was conducted under the oversight of a Johns Hopkins institutional review board (IRB00228485 and IRB00268645). This board follows the principles of the Declaration of Helsinki. Informed consent was waived because the source of the data was the Johns Hopkins Electronic Medical Record, which contains patient information collected as part of routine medical care. It would have been impossible to collect this information on such a large number of patients without unduly interrupting the clinical workflow of data capture without a waiver. All downstream analyses using this resource were done using Health Insurance Portability and Accountability Act (HIPAA) defined limited datasets.

