## Supplemental for "Real-world observation on response to cholinesterase inhibitors or selective serotonin reuptake inhibitors prescribed to outpatients with dementia using electronic medical records"

A) SSRI medication usage

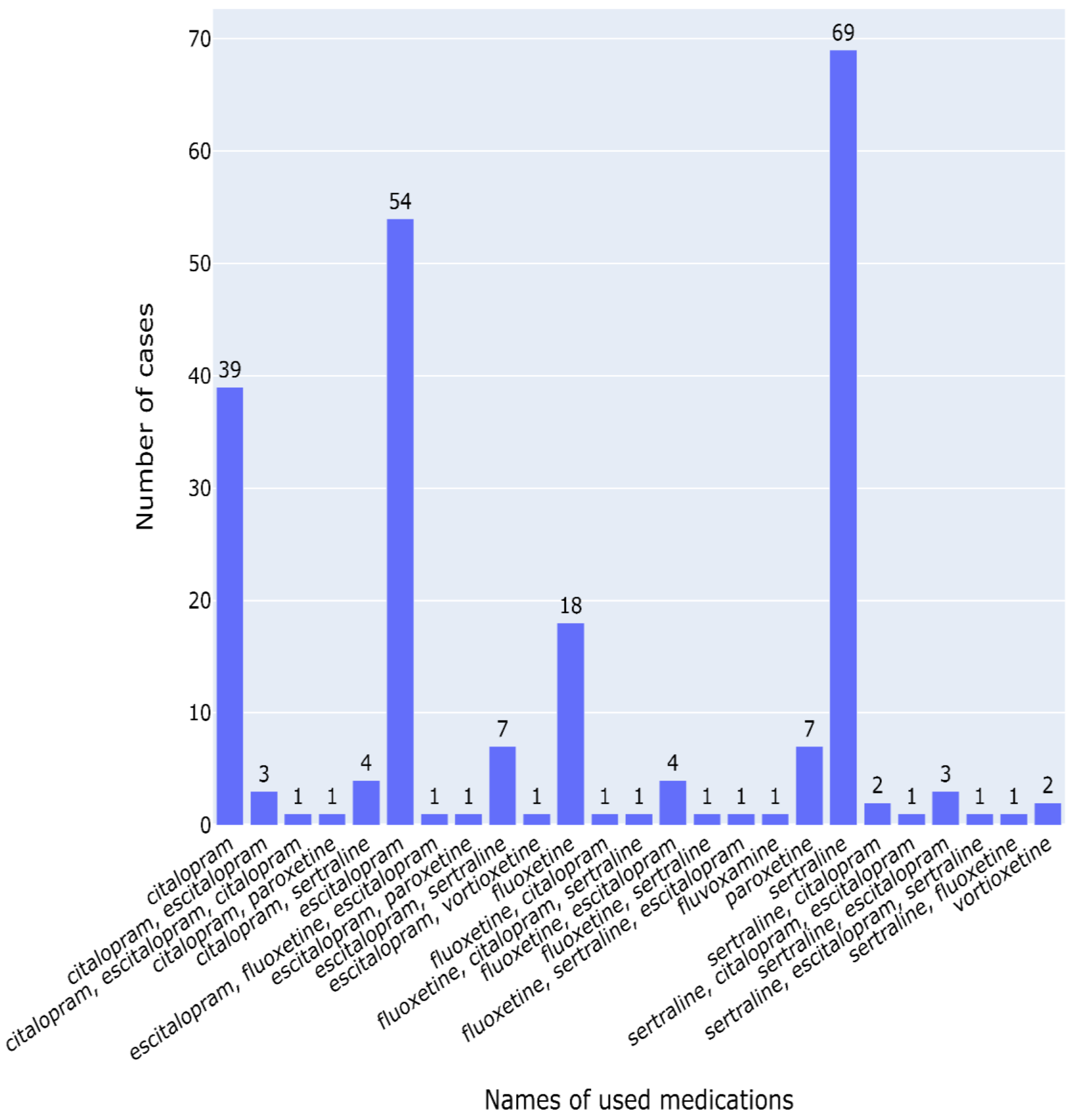

B) CEI medication usage

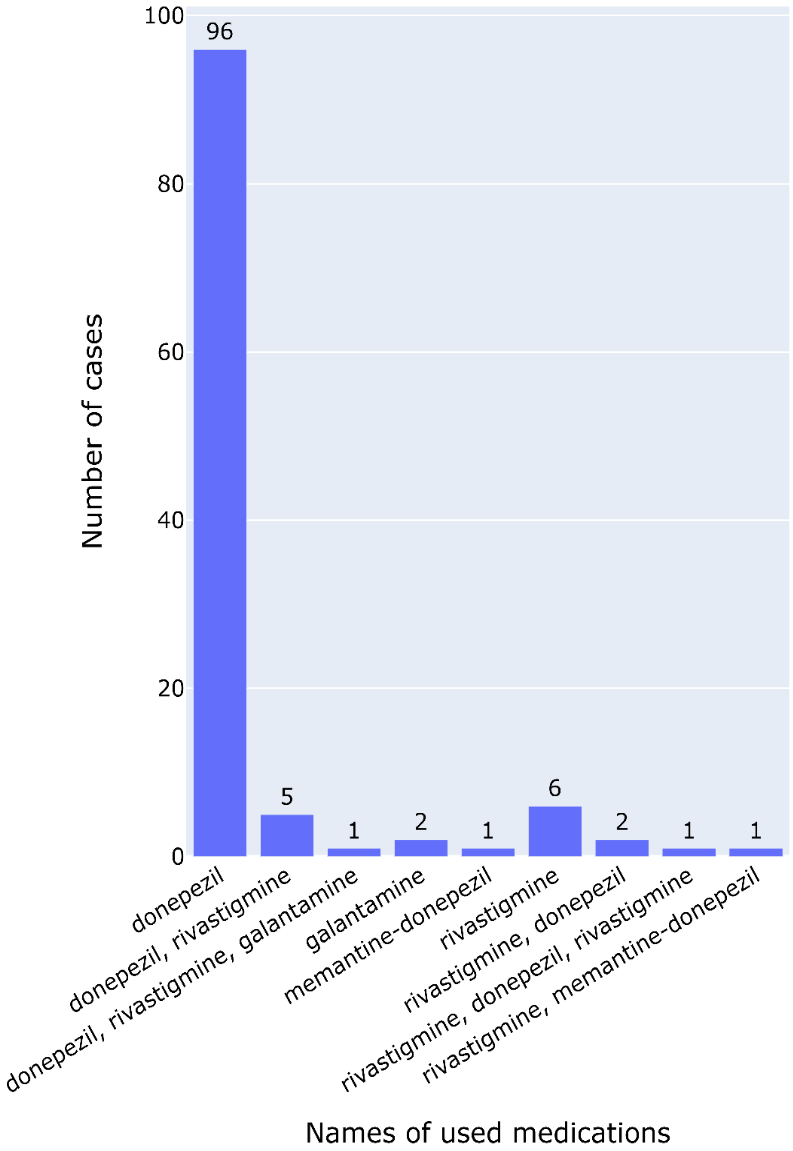


**Supplemental Figure-1.** The lists of medications used for SSRI and CEI cases. Medications names that have more than 2 drugs are switched within 3-months washout period and separated by commas in the order of usage.

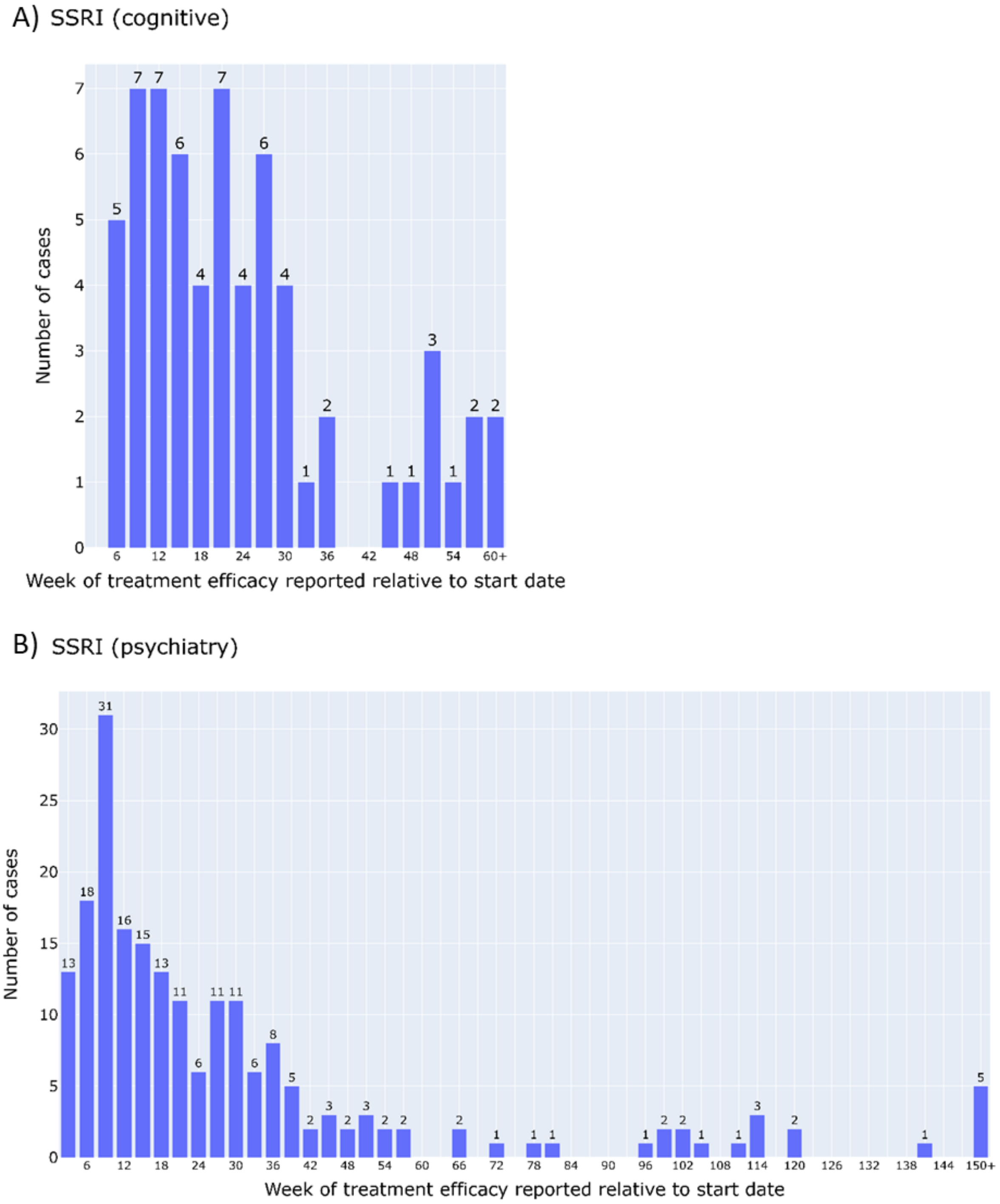
A) SSRI (cognitive)
Number of cases
0 1 2 3 4 5 6 7
6 12 18 24 30 36 42 48 54 60+
Week of treatment efficacy reported relative to start date
B) SSRI (psychiatry)
Number of cases
0 5 10 15 20 25 30
6 12 18 24 30 36 42 48 54 60 66 72 78 84 90 96 102 108 114 120 126 132 138 144 150+
Week of treatment efficacy reported relative to start date

C) CEI (cognitive)

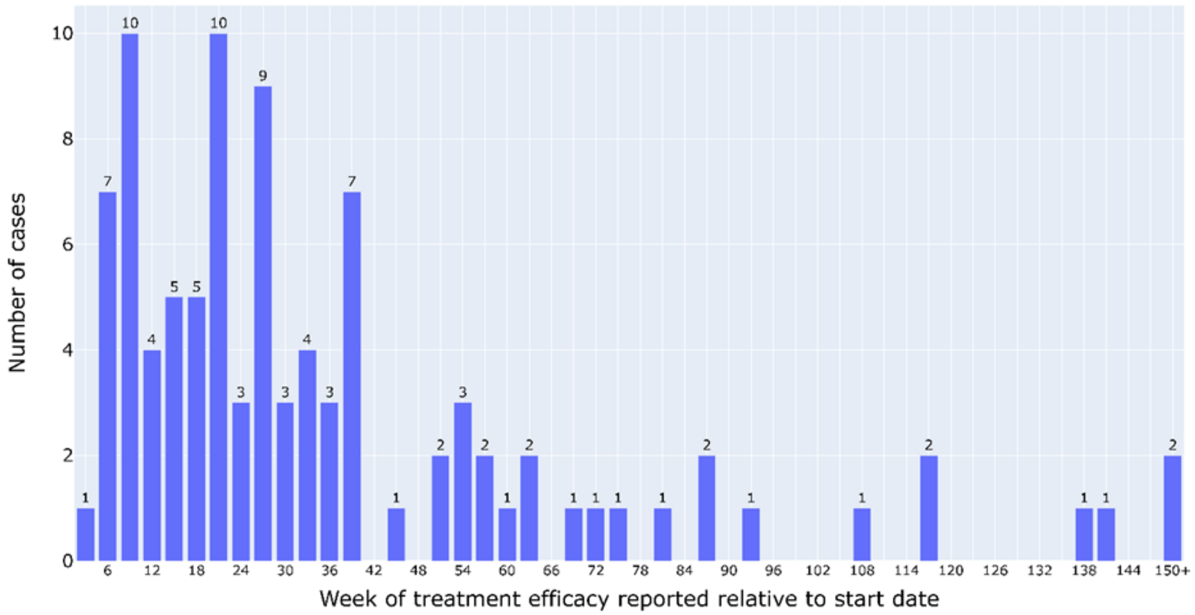


D) CEI (psychiatry)

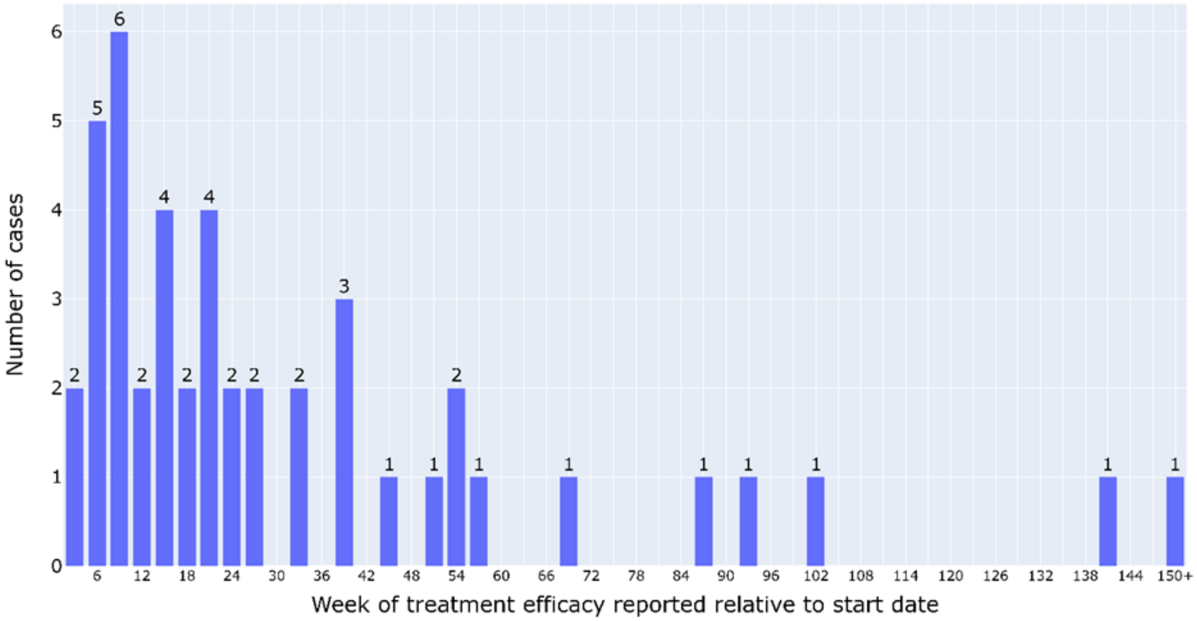


**Supplemental Figure-2.** Time of medication start to determination of treatment efficacy

**Supplemental Table-1.** Krippendorff’s alpha reliability test

| | | Cognitive NOTE | Psychiatric NOTE | Cognitive CIBIC-plus | Psychiatric CIBIC-plus |
|---|---|---|---|---|---|
| ICC (type=ICC2k) based on ordinal assumption (worsen < no change < improved) excluding N/A values | coefficient | 1 | 1 | 1 | 1 |
| | *P*-value | <0.001 | <0.001 | <0.001 | <0.001 |
| | number of subjects | 7 | 10 | 5 | 4 |
| Krippendorff's alpha | alpha | 0.779 | 1 | 1 | 1 |
| | number of subjects | 12 | 12 | 12 | 12 |
| Dice coefficient | N/A | 0.75 | 1 | | |
| | improve | 1 | 1 | | |
| | no change | 1 | 1 | | |
| | worse | 0.75 | 1 | | |
